# Impact of SARS-CoV-2 Infection on Antimicrobial Resistance Gene Profiles in the Upper Respiratory Tract: A Cross-Sectional Study

**DOI:** 10.1101/2024.11.14.24317312

**Authors:** Siddharth Singh Tomar, Krishna Khairnar

## Abstract

**Objectives:** To investigate the impact of SARS-CoV-2 infection on the antimicrobial resistance (AMR) gene profiles in the upper respiratory tract (URT) and To evaluate variations in AMR gene diversity, abundance, and ESKAPE-associated AMR in URT. By comparing SARS-CoV-2-positive patients to healthy controls.

**Methods:** 95 URT swab samples from SARS-CoV-2-positive (n=48) and RTPCR-negative control participants (n=47) collected from central India. Metagenomic DNA was extracted, and metagenomic sequencing was performed using the Illumina NextSeq550 platform. Sequencing data were analysed using the Chan Zuckerberg ID pipeline for Antimicrobial resistance (AMR) gene detection and taxonomic profiling. Chao1, Shannon and Simpson diversity indices, Bray-Curtis dissimilarity, and Bayesian regression, were used to identify significant differences in AMR gene abundance and microbial associations.

**Results:** The Chao1 index (p=0.01651) of SARS-CoV-2 samples indicated significantly higher AMR gene richness than the controls. Resistance genes, such as mecA, blaOXA-48, and blaNDM-1, showed higher abundance in SARS-CoV-2 samples. These genes were found to be linked to high-priority pathogens like *Klebsiella pneumoniae, Escherichia coli,* and *Staphylococcus aureus*. Bayesian regression demonstrated that SARS-CoV-2 infection is a significant factor in elevated AMR gene abundance (β = 1.549, HDI [1.409, 1.691]). Females showed higher AMR levels than males (β = 0.261, HDI [0.167, 0.350]), and the model outputs showed no significant age correlation. Sankey diagrams and heatmaps showed higher AMR gene diversity and abundance in the SARS-CoV-2 group.

**Conclusions:** SARS-CoV-2 infection alters the URT’s AMR gene profile and increases the resistance genes’ abundance and diversity. The results indicate a requirement to enhance AMR surveillance of COVID-19 patients to adapt antimicrobial stewardship strategies and reduce the chances of secondary infections. It is, therefore, essential to carry out more extensive studies to analyze temporal variations and the effects of antibiotic overuse on AMR evolution.

## Introduction

The global SARS-CoV-2 pandemic has induced research into its broader effects. This study focuses on the Antimicrobial resistance (AMR) dynamics in the upper respiratory tract (URT) of SARS-CoV-2-infected individuals. The URT is the primary entry point and predisposition site for respiratory viruses. Therefore, URT is an important site for studying AMR diversity in SARS-CoV-2-infected individuals. SARS-CoV-2 infection may contribute to dysbiosis of baseline URT flora and promote the spread of AMR genes¹, ²,³. Respiratory viral infections have been shown to alter microbial diversity and contribute to secondary bacterial infectionsL and favoring pathogenic bacteria capable of acquiring resistanceL.

AMR in respiratory infections is a growing concern due to widespread antibiotic use even before the SARS-CoV-2 pandemic. Indiscriminate antibiotic use disrupts the microbial balance and promotes the enrichment of resistant organismsL. Respiratory resistomes serve as reservoirs for resistance genes transferable to pathogenic bacteriaL. Studies have shown that SARS-CoV-2 infection influences microbial communities and resistomes of patientsL. Virus-induced inflammation and immune suppression promote bacterial growth and enrichment of AMR genes in SARS-CoV-2-positive individuals. Recently, the World Health Organisation has also highlighted the overuse of antibiotics in COVID-19 patients contributing to increasing AMR¹L.

Microbiome studies on COVID-19 patients reveal shifts in diversity and increased abundance of clinically important pathogens like *Staphylococcus aureus* and *Pseudomonas aeruginosa*. These pathogens can acquire resistance to common antibiotics¹¹. The AMR genes like blaKPC, blaNDM, and mecA in COVID-19 patients, responsible for resistance in ESKAPE pathogens, were detected in the resistomes of SARS-CoV-2 positive individuals^12^. Despite growing evidence of SARS-CoV-2’s impact on the resistome, further research is required to fully understand the effects of viral infection on AMR abundance and diversity, for which metagenomic sequencing is a valuable tool¹³.

Few studies have investigated the relationship between SARS-CoV-2 infection and changes in the URT resistome in the Indian context. Researchers investigated changes in the URT microbiome of SARS-CoV-2-infected individuals to identify infection-specific signatures for developing nasal prebiotic therapies¹L. However, they did not examine AMR alterations in the URT of SARS-CoV-2-positive individuals. This study uses a metagenomic approach to understand the effect of SARS-CoV-2 infection on URT resistome and abundance of ESKAPE pathogens.

## 2. Materials and methods

### 2.1. Study Population

The samples used for the study were collected during March-April 2023. The study population belongs to the Vidarbha region of Central India, with samples collected from participants across five districts: Nagpur, Wardha, Gadchiroli, Chandrapur, and Bhandara. For the SARS-CoV-2 Negative Group (Control), the median age (interquartile range [IQR]) is 29 years (22 to 37). For the SARS-CoV-2 Positive Group, the median age (IQR) is 36 years (19 to 67). Participants from the SARS-CoV-2 group presented with severe acute respiratory infection (SARI) and/or influenza-like illness (ILI) symptoms. While the participants from the control group were asymptomatic and RTPCR negative.

### 2.2. Sample Collection and Processing

In total, 96 URT swab samples in Viral Transport Medium (VTM) were collected for this study, out of which 48 were from SARS-CoV-2 positive individuals (SARS-CoV-2 group) and 48 samples were collected from Healthy control (RTPCR negative). These samples were collected by expert healthcare professionals while following the standard sample collection guidelines. The collected samples were maintained at 4 °C ≤ 5 days (Short-term storage) and at -80 °C for the long term. Aliquots of these collected samples were then processed for SARS-CoV-2 RTPCR testing under Biosafety level-II conditions. Samples with ≤ 25 cycle threshold value of SARS-CoV-2 target genes were considered positive samples.

### 2.3. Metagenomic DNA Extraction, Library Preparation, and Metagenomic Sequencing

The metagenomic DNA extraction was done using the QIAamp DNA Microbiome Kit (Catalog No. 51704). The DNA concentration was measured using a Qubit fluorometer. DNA purity was assessed using a Nanodrop spectrophotometer by analyzing the A260/280 and A260/230 ratios. DNA libraries were prepared with the QIAseq FX DNA Library Preparation Kit. Metagenomic next-generation sequencing (mNGS) was performed on the NextSeq550 platform using a 2x150 bp high-output kit, generating paired-end reads over 300 cycles.

### 2.4. Metagenomic Data Analysis

Metagenomic data analysis was performed using the Chan Zuckerberg ID (CZID) web-based platform. Sequencing quality control was done by removing External RNA Controls Consortium (ERCC) sequences with Bowtie2 (Version 2.5.4)^16^. Sequencing adapters, short reads, low-quality sequences, and low-complexity regions were filtered using a customized fastp tool^17^. DNA sequences with quality scores < 17, reads shorter than 35 bp, high-complexity sequences > 40%, and > 15 undetermined bases were excluded.

The Host (Human) sequences were removed by aligning with Bowtie2 and HISAT2 against reference genomes^18^. CZID-dedup was used for 100% identical sequences till the first 70 base pairs. Only one representative read was retained. The STAR algorithm^19^ was also used to remove duplicate, low-quality, and low-complexity host reads. Non-human reads were aligned to the NCBI nucleotide and protein databases (NCBI Index Date: 06-02-2024) using GSNAPL and RAPSearch. Post-filtering, sequences were aligned to the NCBI nucleotide (NT) database with Minimap2^20^ and the NCBI protein (NR) database with Diamond^21^. Hits were annotated with accession numbers, and taxon counts were generated from GSNAP and RAPSearch results.

SPAdes was used for de novo (without reference) genome assembly. The original reads were mapped to assembled contigs with Bowtie2. BLAST analysis was then conducted on the contigs against the Nucleotide NT-BLAST database (GSNAP) and the Protein NR database (RAPSearch2).

### 2.5. AMR pipeline

For AMR analysis CZID’s AMR Pipeline v1.4.2 ^22^ was used. AMR gene detection is performed using the Resistance Gene Identifier (RGI) tool. The Comprehensive Antibiotic Resistance Database (CARD) v3.2.6 ^23^ was used to identify AMR genes and pathogen species. Contigs of AMR genes are assembled using SPAdes and aligned with CARD through BLAST for species identification. The result files generated from this pipeline contain detailed information on AMR genes, quality control metrics, and pathogen-of-origin detection.

### 2.6. Statistical Analysis and Visualization

Python 3.10.12 was used for statistical analysis and visualization. The analysis used the following Python packages: SciPy (version 1.13.1) for t-tests and ANOVA. statsmodels (version 0.14.4) for regression models and hypothesis testing. scikit-learn (version 1.5.2) was used for PCA, PCoA, Multiple regression and additional statistical analysis. For data visualization, matplotlib (version 3.7.1) was used. Seaborn (version 0.13.2) was used for statistical visualizations with heatmaps and violin plots. Interactive visualizations like the Sankey diagram were created using Plotly (version 5.24.1). Data transformation tasks such as pivoting and log transformation were handled with Pandas (version 2.2.2). Statistical analysis was performed using numpy (version 1.26.4). The abundance metric of reads per million (RPM) was used for statistical analysis and data visualisation.

## 3. Results

### 3.1. Summary of Sequencing Results

Metagenomic sequencing of SARS-CoV-2 (48 samples) and Control (47 samples) groups was done using the Illumina NextSeq550 platform. The SARS-CoV-2 group generated an average of 7.26 ± 1.52 million reads, out of which 25% (1.78 ± 0.56 million reads) passed the human filter. The control group had generated average 6.52 ± 3.47 million reads out of which 15% (0.78 ± 0.37 million reads) passed the human filter. One sample from the control group was not included in the study due to quality check failure before sequencing.

### 3.2. AMR Gene Alpha Diversity Analysis

The violin plots **(Figure 1)** show that the SARS-CoV-2 group has significantly higher species richness than the control group. This is reflected by the Chao diversity index (p = 0.01651), implying that the SARS-CoV-2 group harbors more distinct taxa. However, the Shannon and Simpson diversity indices show no significant differences between the groups (p = 0.98515 and p = 0.90225, respectively). This indicates that while the SARS-CoV-2 group has greater species richness, the overall distribution and balance of species abundances are similar to those in the control group.

**Figure 1:**
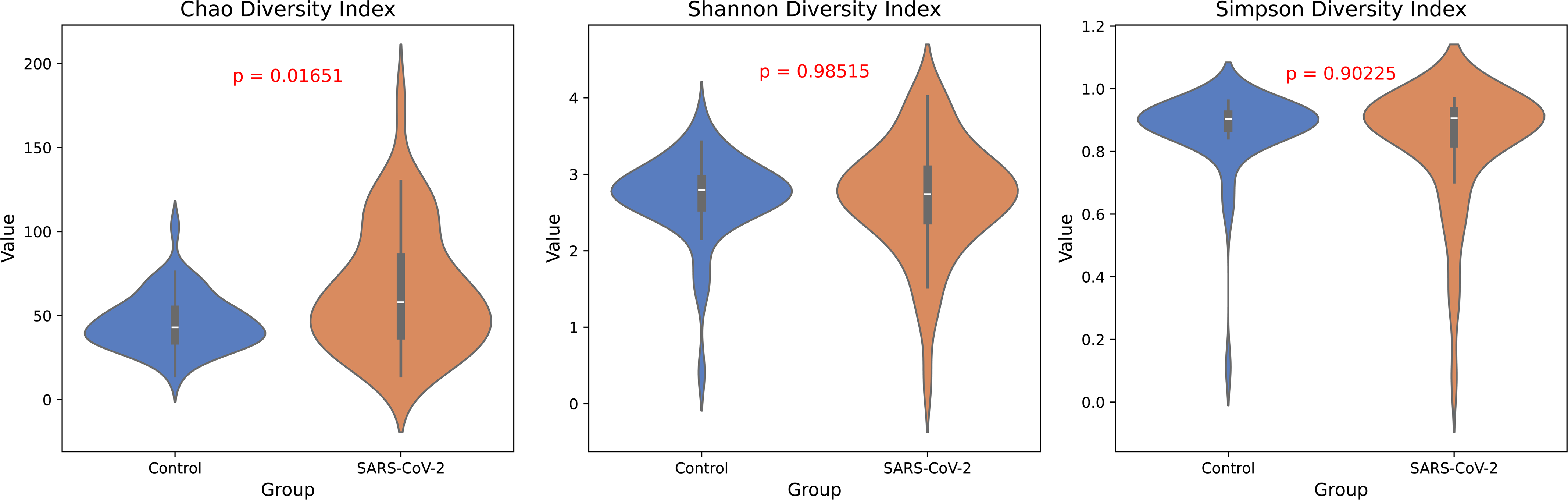
Chao1, Shannon, and Simpson Diversity of AMR Genes: Chao1 diversity (left), Shannon diversity (center), and Simpson diversity (right). Chao1 diversity (p=0.01651), Shannon (p = 0.98515), and Simpson (p = 0.90225)

### 3.3. AMR Gene Beta Diversity: Bray-Curtis Dissimilarity and PCoA Analysis

The Bray-Curtis dissimilarity heatmap **(Figure 2)** shows distinct clustering of SARS-CoV-2 (red sample tags) and control (blue sample tags) samples. Significant within-group similarity in both groups was observed, as indicated by lighter shades. The higher dissimilarity between groups is seen in the off-diagonal regions. The diagonal line represents zero dissimilarity. The Bray-Curtis dissimilarity heatmap highlights clear within-group similarities and significant between-group differences in AMR profiles.

**Figure 2:**
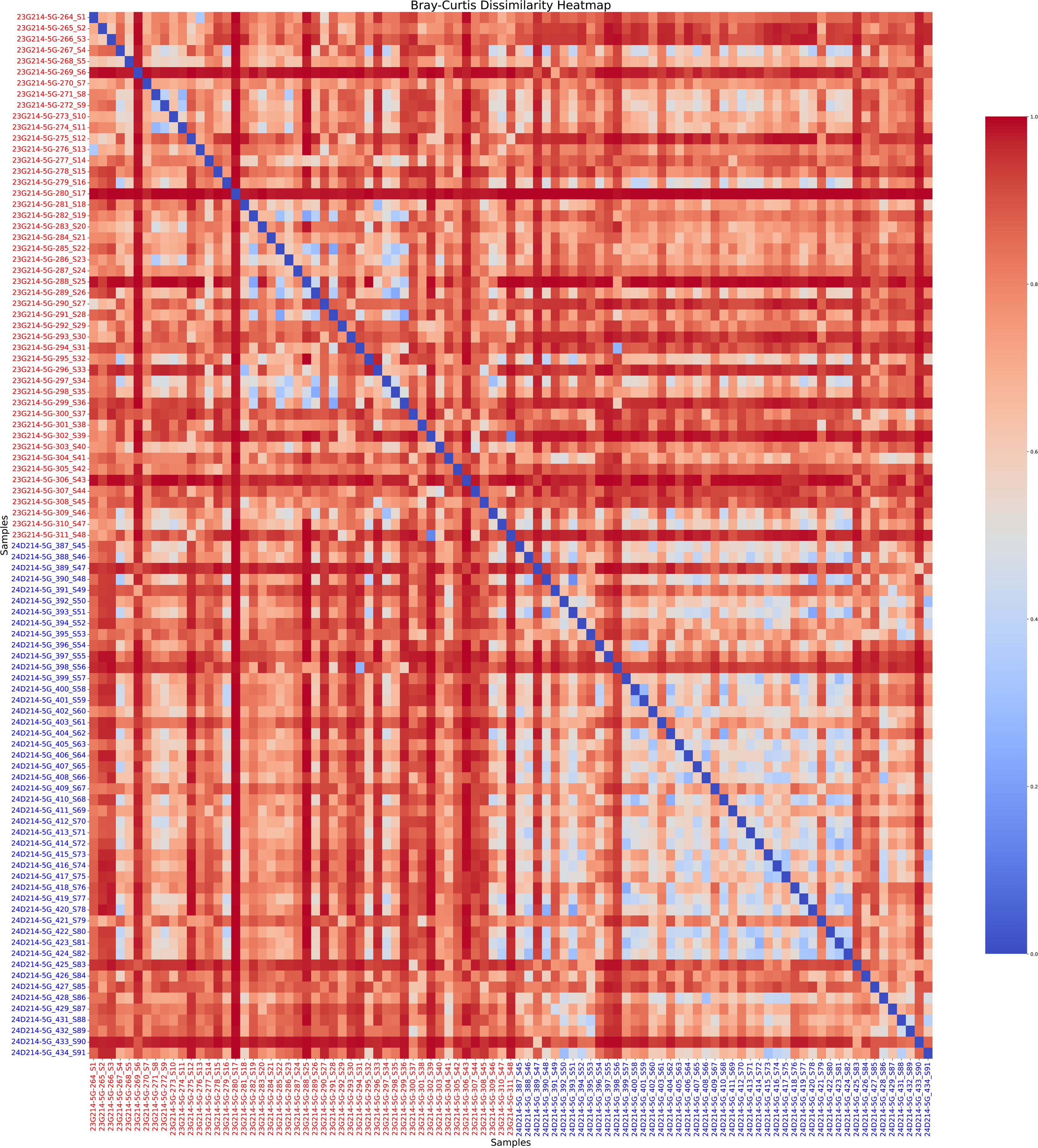
Bray-Curtis Dissimilarity Heatmap: The heatmap shows SARS-CoV-2 samples (Sample IDs in red) and control samples (Sample IDs in blue). Red shades of heatmap represent dissimilarity, while blue represents similarity. The diagonal represents perfect similarity (self-comparison), indicated in dark blue.

**(Figure 3)** represents PCA (left) and PCoA (right) plots comparing SARS-CoV-2 (red) and Control (blue) groups. In the PCA plot, PC1 (77.90% variance) and PC2 (7.91% variance) reveal that the control group is tightly clustered near the origin. While, the SARS-CoV-2 group shows a greater spread along PC1. In the PCoA plot, Axis 1 (32.79% variance) and Axis 2 (13.42% variance) show the control group forming a compact cluster. In contrast, the points in the SARS-CoV-2 group are more scattered. The groups are clearly distinct along Axis 1 in both plots, with SARS-CoV-2 exhibiting higher variations.

**Figure 3:**
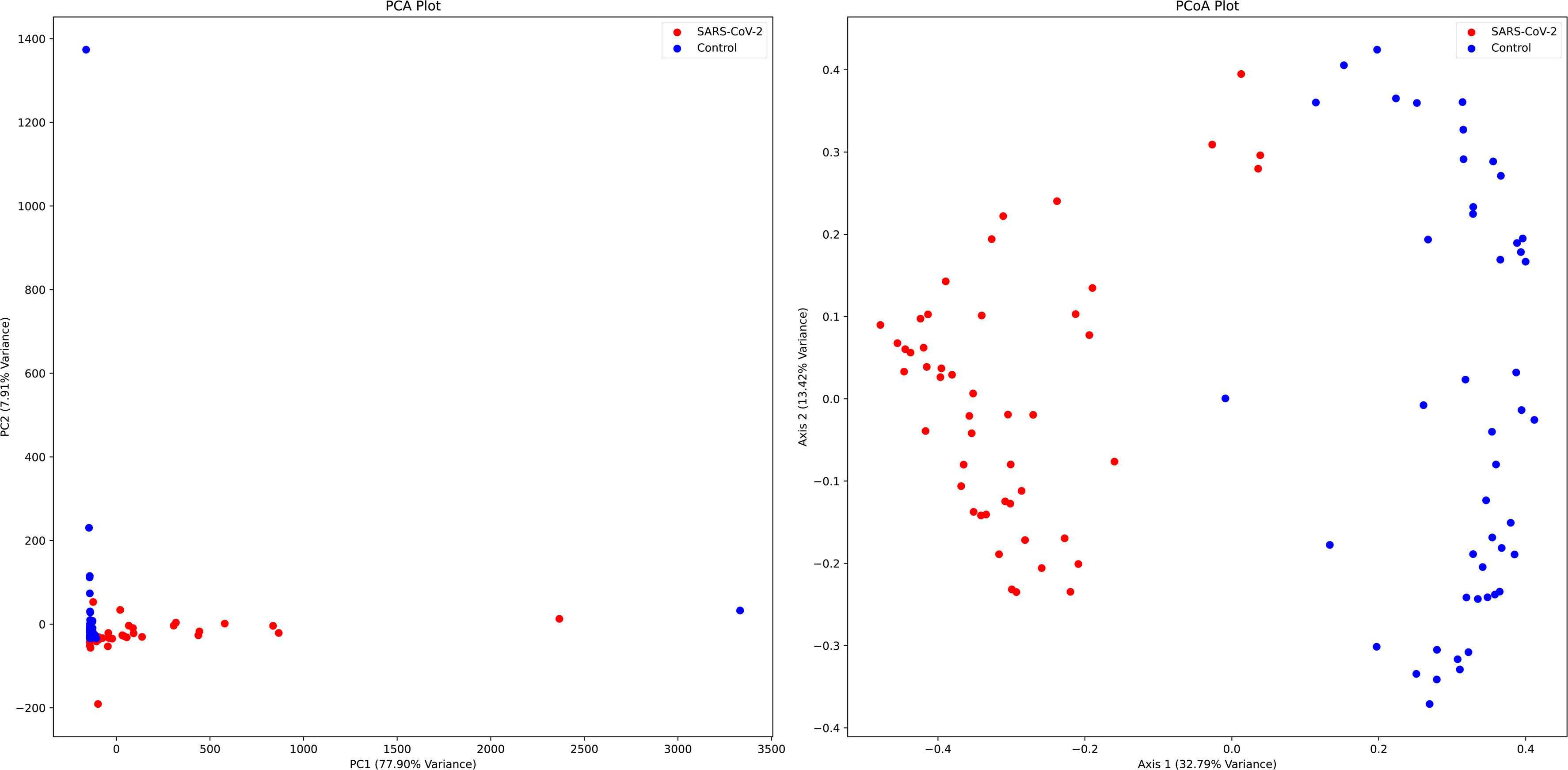
PCA (left) and PCoA (right) plots showing the clustering of AMR gene profiles in control and SARS-CoV-2 groups. PCA (PC1: 77.90% variance) and PCoA (Axis 1: 32.79% variance) both show distinct separation between the groups.

### 3.4. Differences in Resistome and Pathogen profiles among SARS-CoV-2 and Control Groups

Antimicrobial resistance (AMR) gene abundance and pathogen profiles were significantly differing among the SARS-CoV-2 and control groups. **(Figure 4)** presents a heatmap of statistically significant (p < 0.05, Mann-Whitney) AMR genes. It was observed that the SARS-CoV-2 group exhibited a higher density and more consistent abundance of genes such as *mecA, blaOXA-48*, and *blaNDM-1*. Meanwhile, the control group showed sparse and less pronounced patterns of AMR gene abundance.

**Figure 4:**
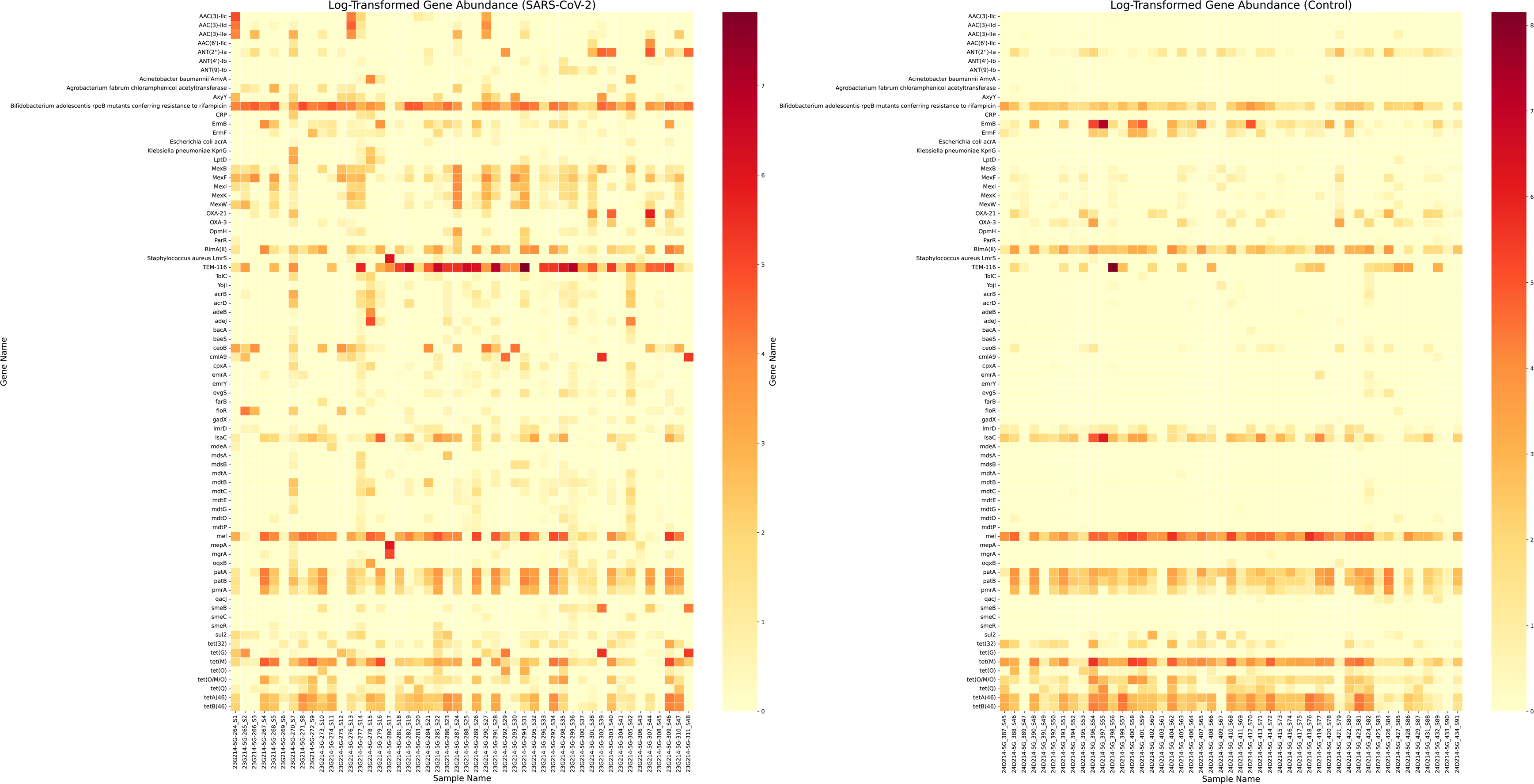
Compositional Variations in the abundance of AMR Genes Among Control and SARS-CoV-2 Groups (Log-Transformed Significant Genes). Each row represents a distinct AMR gene, and each column represents a sample. The color intensity reflects gene abundance, with darker colors indicating higher abundance.

**(Figure 5)** shows species-specific resistome profiles using a heatmap visualization. Significant associations between AMR genes and microbial species were observed in the SARS-CoV-2 group. Higher abundances of resistance genes were found in species such as *Klebsiella pneumoniae, Escherichia coli*, and *Staphylococcus aureus*. The control group exhibited weaker associations and lower resistance gene abundances. The AMR genes in the control group were primarily linked to species like *Acinetobacter baumannii* and *Pseudomonas aeruginosa*.

**Figure 5:**
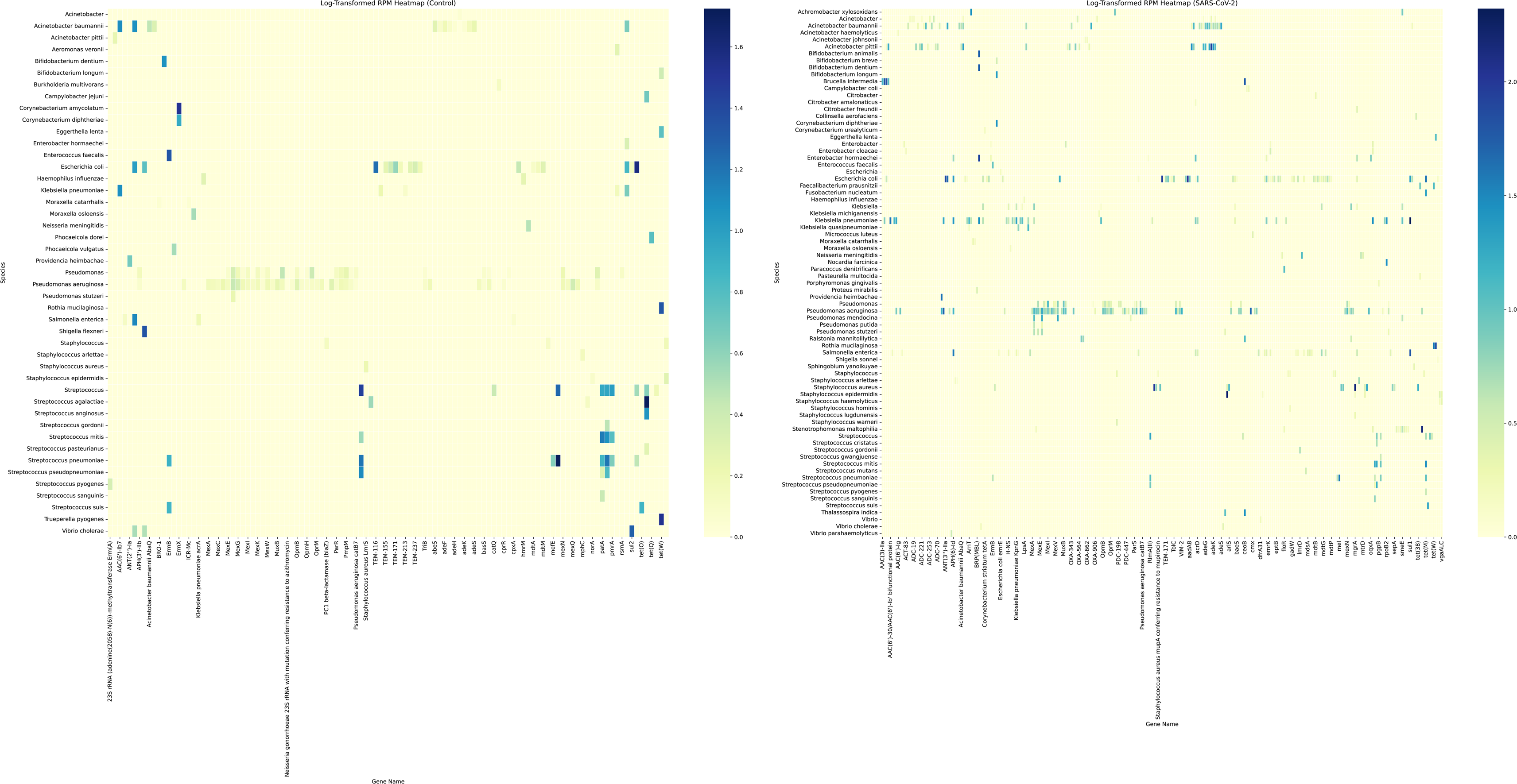
Variations in the Abundance of Species-associated AMR Genes Among Control and SARS-CoV-2 Groups (Log-Transformed) The heatmaps display log-transformed read counts per million (RPM).

The Sankey diagram **(Figure 6)** showed the distribution of AMR genes within high-priority ESKAPE pathogens and their relative abundances among sample groups. Each ribbon represents an AMR gene, with the width proportional to its abundance (RPM). This reveals distinct differences in the distribution of AMR genes between the two groups. The SARS-CoV-2 group showing a larger diversity and higher abundance of ESKAPE-associated AMR genes.

**Figure 6:**
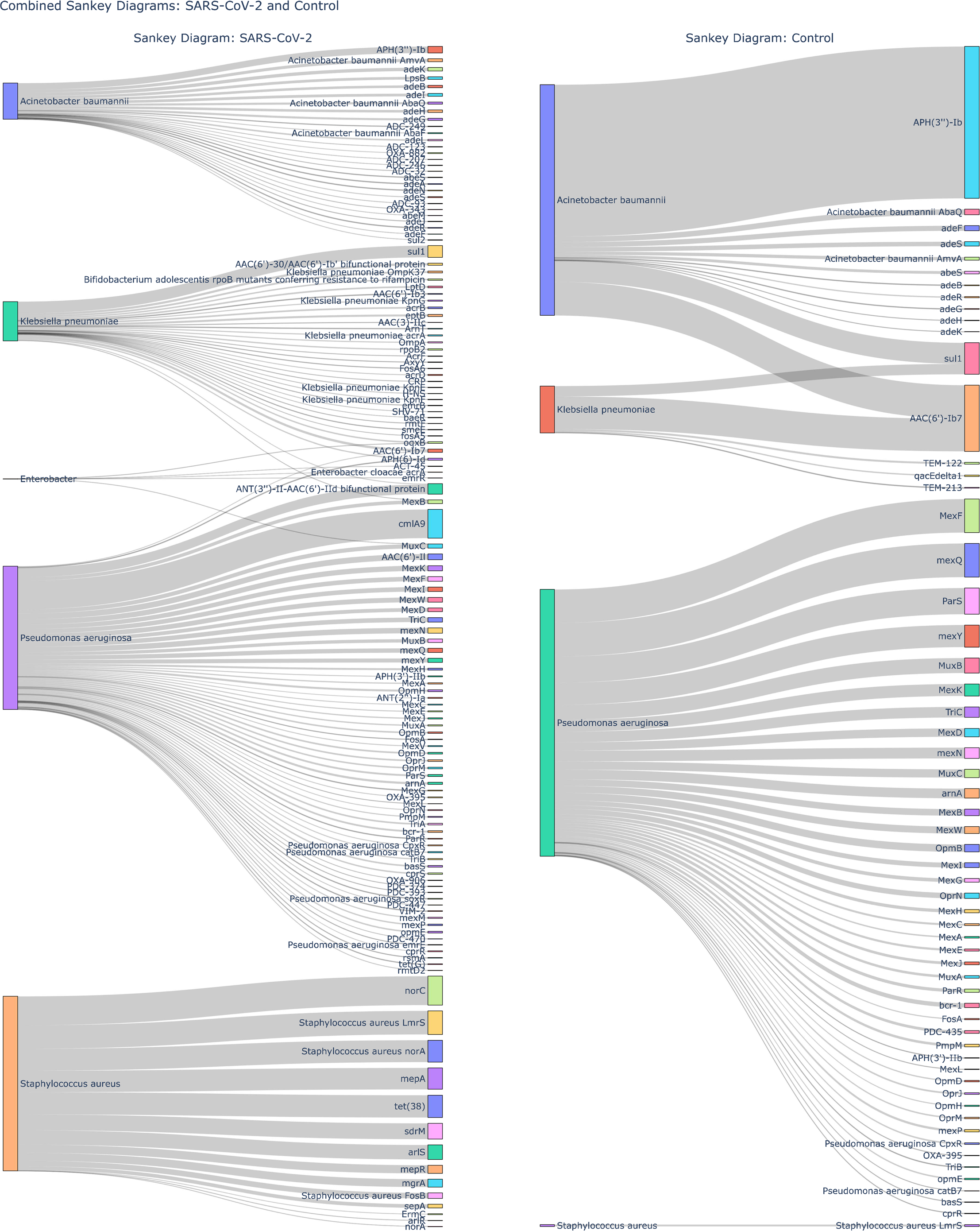
Sankey Diagram Showing the Relationship Between High-Priority ESKAPE Pathogens and AMR Genes in SARS-CoV-2 and Control Groups. The width of the flows between species and AMR genes indicates the abundance in RPM.

### 3.5. Differential Abundance of AMR Genes

The volcano plot **(Figure 7)** displays the differential abundance of AMR (antimicrobial resistance) genes between the SARS-CoV-2 and Control groups. This representation is based on log2 fold change on (x-axis) and statistical significance (-log10 p-value), on y-axis). Genes with log2 fold change > 1 and Mann-Whitney p-values < 0.05 are highlighted in red as significant. Dashed lines in the figure represent thresholds for significance. Horizontal dashed line for the p-value (0.05) and vertical for log2 fold change (±1). The results show that most of the statistically significant genes are more abundant in the SARS-CoV-2 group.

**Figure 7:**
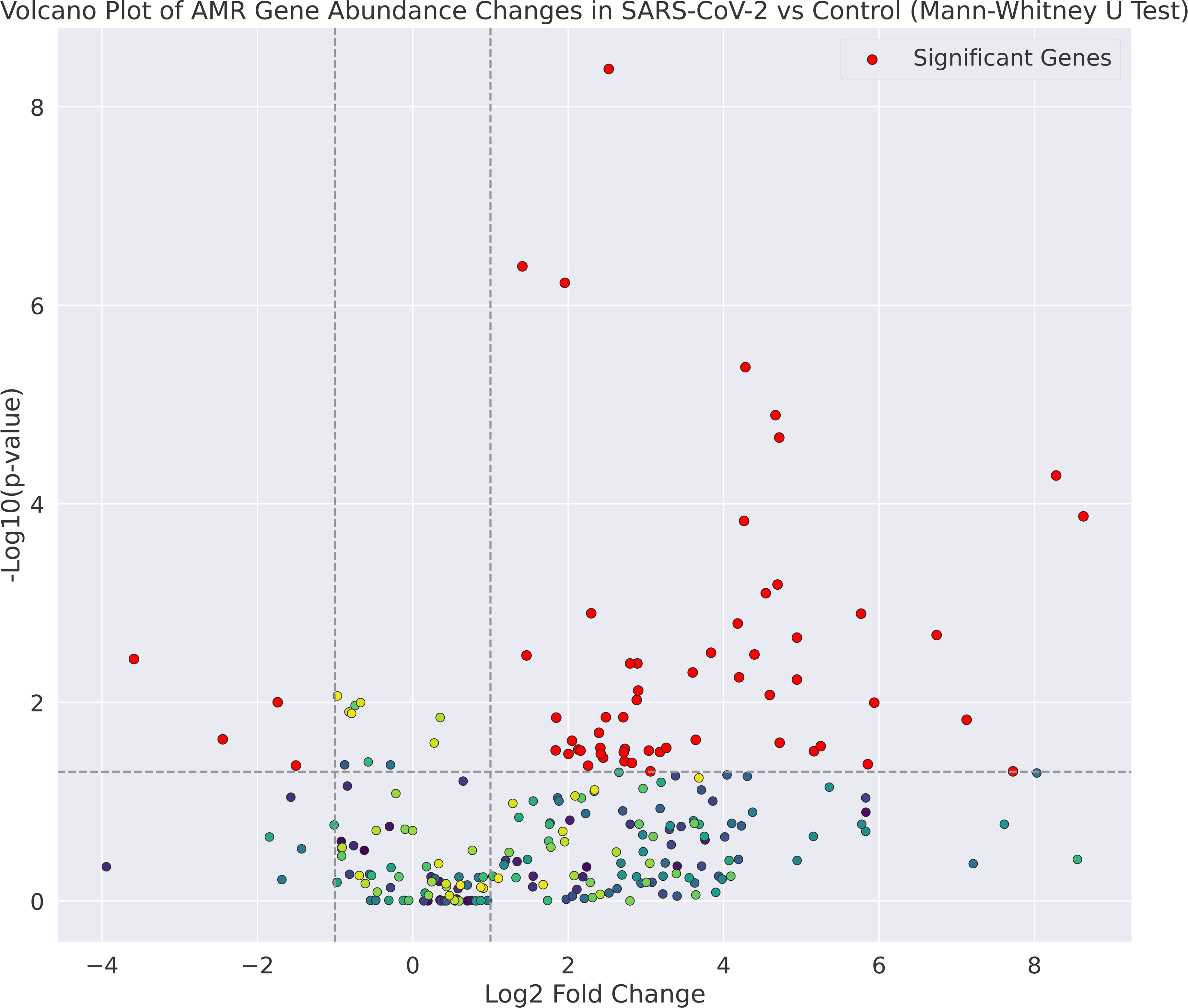
Volcano plot of AMR gene abundance changes between SARS-CoV-2 and control samples, based on a Mann-Whitney U test. The x-axis represents log2 fold change (positive for SARS-CoV-2, negative for control), and the y-axis shows -log10 p-values, with higher values indicating statistical significance. Points above the dashed line at -log10(0.05) are significant (p < 0.05). Gens with |log2 fold change| > 1 and p-value < 0.05 are highlighted in red.

### 3.6. Bayesian Regression Analysis

The Shapiro-Wilk test confirms that the data is not normally distributed in both groups. For Control, the statistic is 0.108 (p = 2.76x10^-49^), and for SARS-CoV-2 data, the statistic is 0.136 with (p 2.74×10^−43^). For Shapiro-Wilk p<0.05 indicates a non-normal distribution of the data. These results align with the skewed distributions seen in the histograms and Q-Q plots **(Figure 8)**. Due to this non-normal distribution of data, the Bayesian regression model was used to examine the factors influencing antimicrobial resistance (AMR) gene abundance. The model has considered variables such as Sample Type (SARS-CoV-2 vs. Control), Collection Location, Host Sex, and Host Age. The model results showed that SARS-CoV-2 samples had significantly higher AMR gene abundance compared to control samples (β = 1.549, HDI [1.409, 1.691]). Females had significantly higher AMR gene abundance (β = 0.261, HDI [0.167, 0.350]) than males. The variable of host age showed no significant effect on AMR gene abundance. The model summary and categorical mappings are elaborated in **(Table 1)** Trace plots **(Figure 9)** showed good parameter mixing and no divergences. The posterior predictive checks confirmed the alignment between model predictions and observed data **(Figure 10)**.

**Figure 8:**
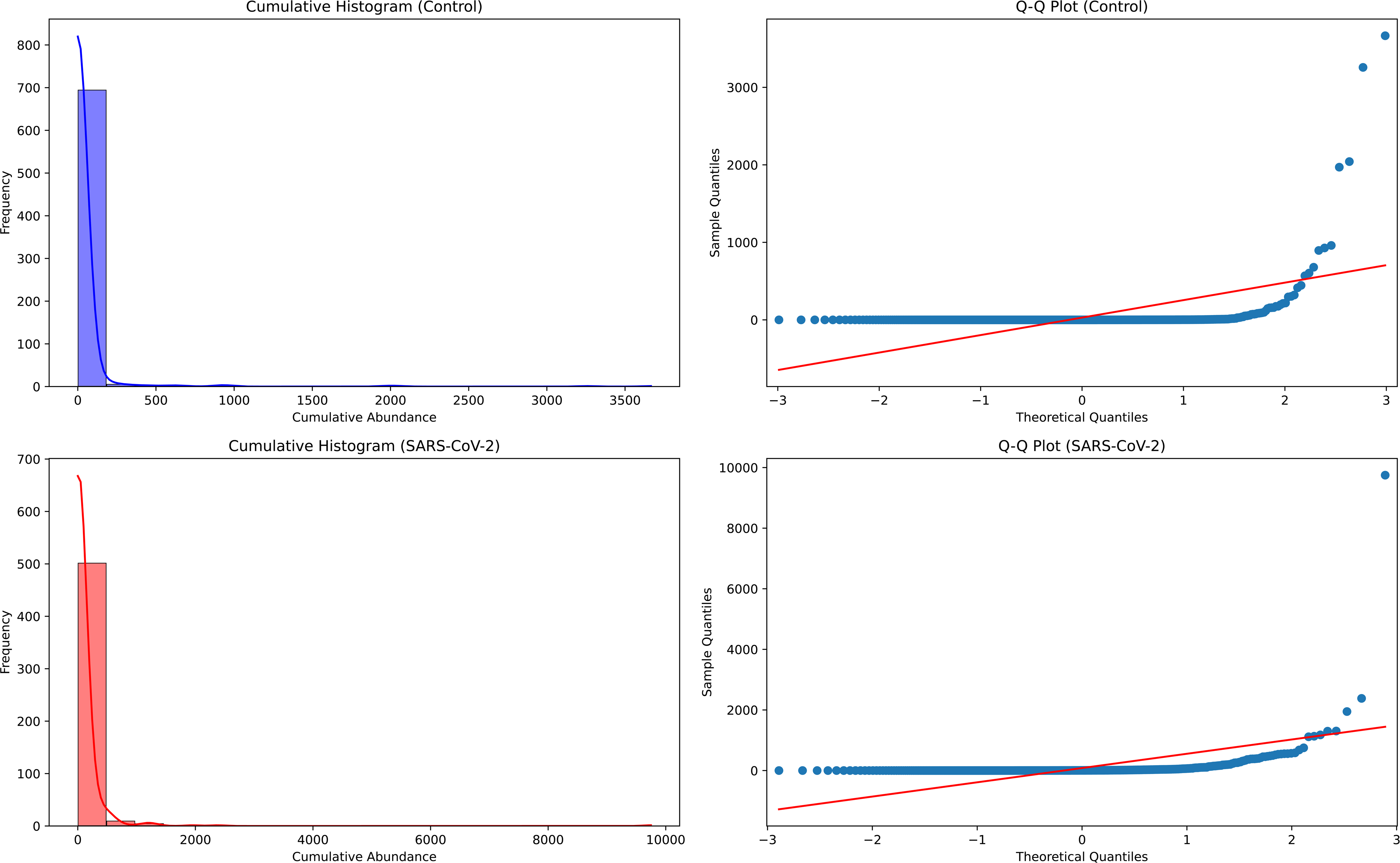
Distributions of antimicrobial resistance gene families in control (top) and SARS-CoV-2 (bottom) groups. Histograms (left). Q-Q plots (right) reveal deviations from normality, with heavier tails in both groups.

**Figure 9:**
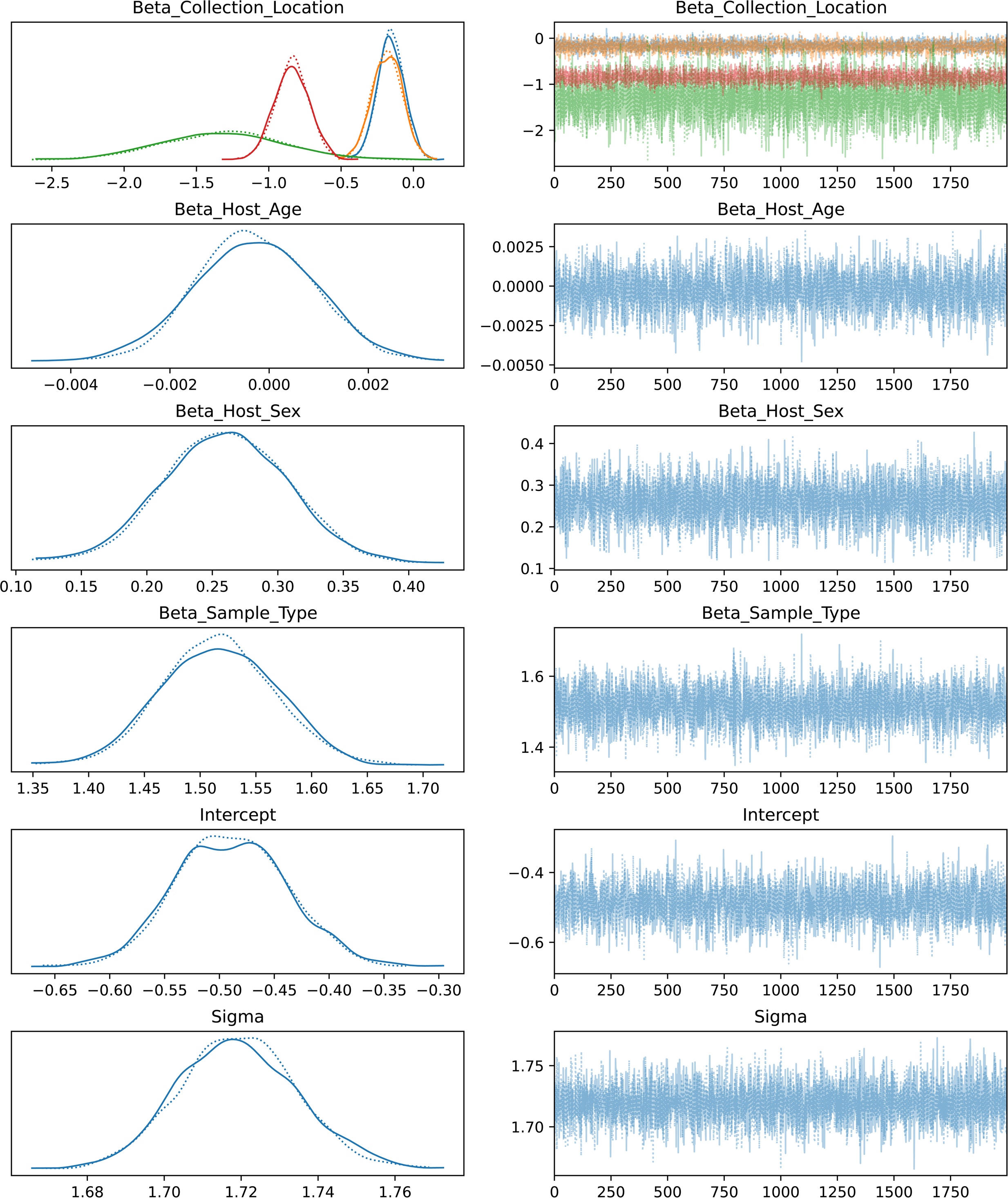
Left panels show posterior density estimates for coefficients: Beta_Collection_Location, Beta_Host_Age, Beta_Host_Sex, Beta_Sample_Type, Intercept, and Sigma. Solid lines indicate posterior densities, while dashed lines represent prior distributions. Right panels display corresponding trace plots of parameter values across iterations, demonstrating adequate mixing and convergence for all parameters.

**Figure 10:**
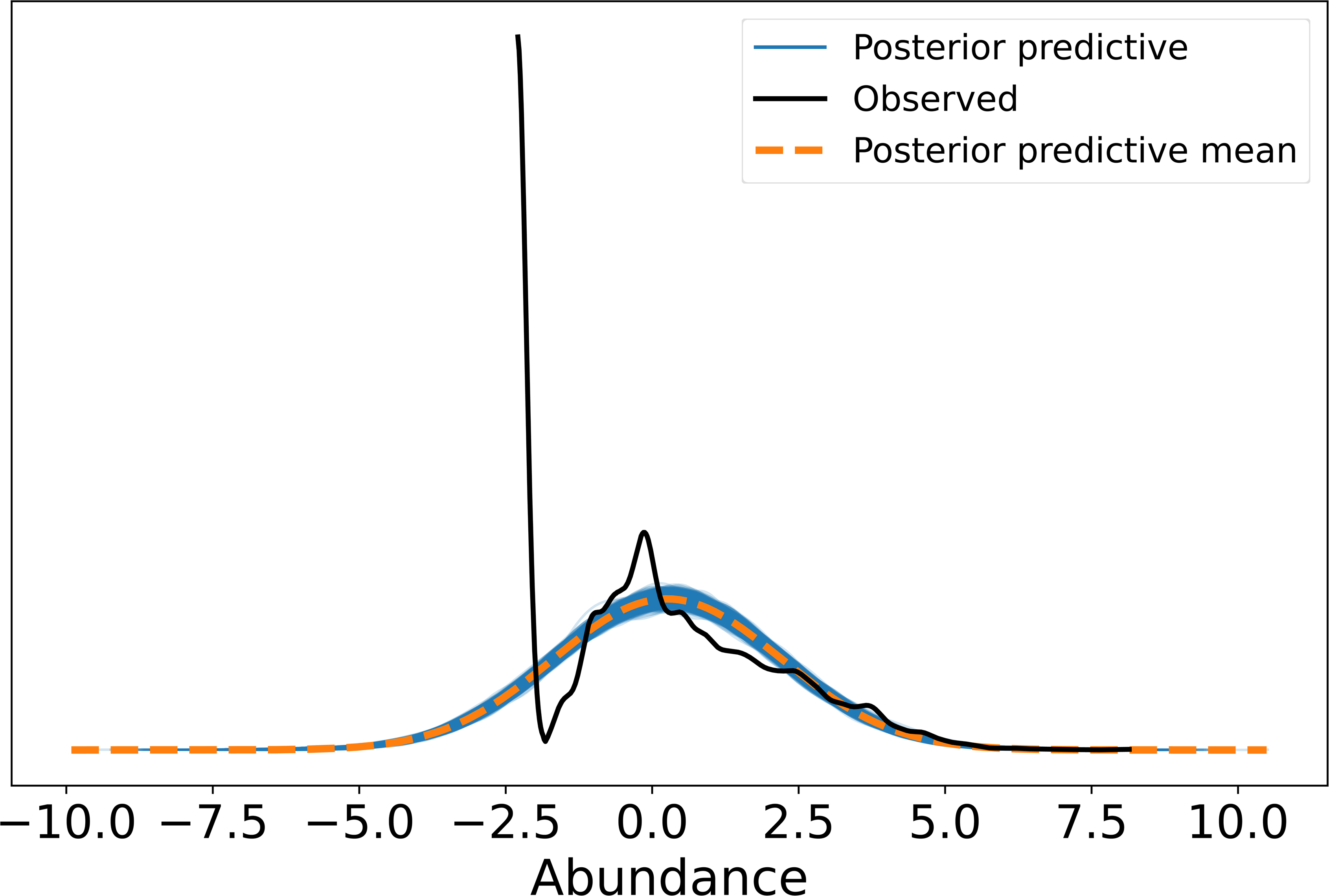
Posterior predictive check comparing observed data (black line) with the posterior predictive distribution (blue line) and its mean (orange dashed line). The alignment of the observed and predicted distributions indicates the model’s ability to capture the underlying data trends.

**Table 1:**
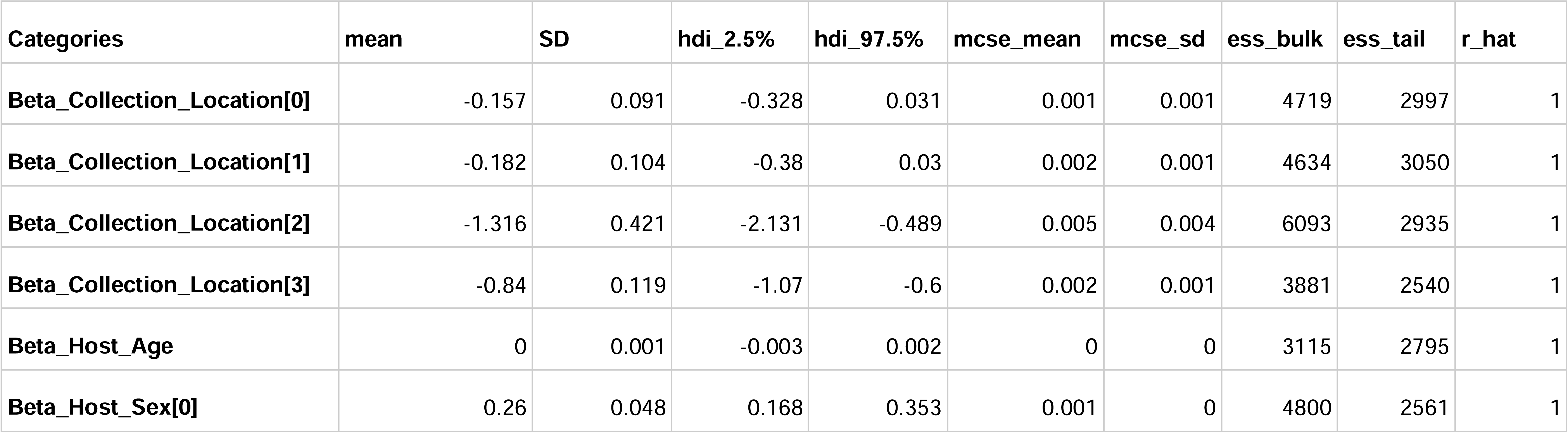
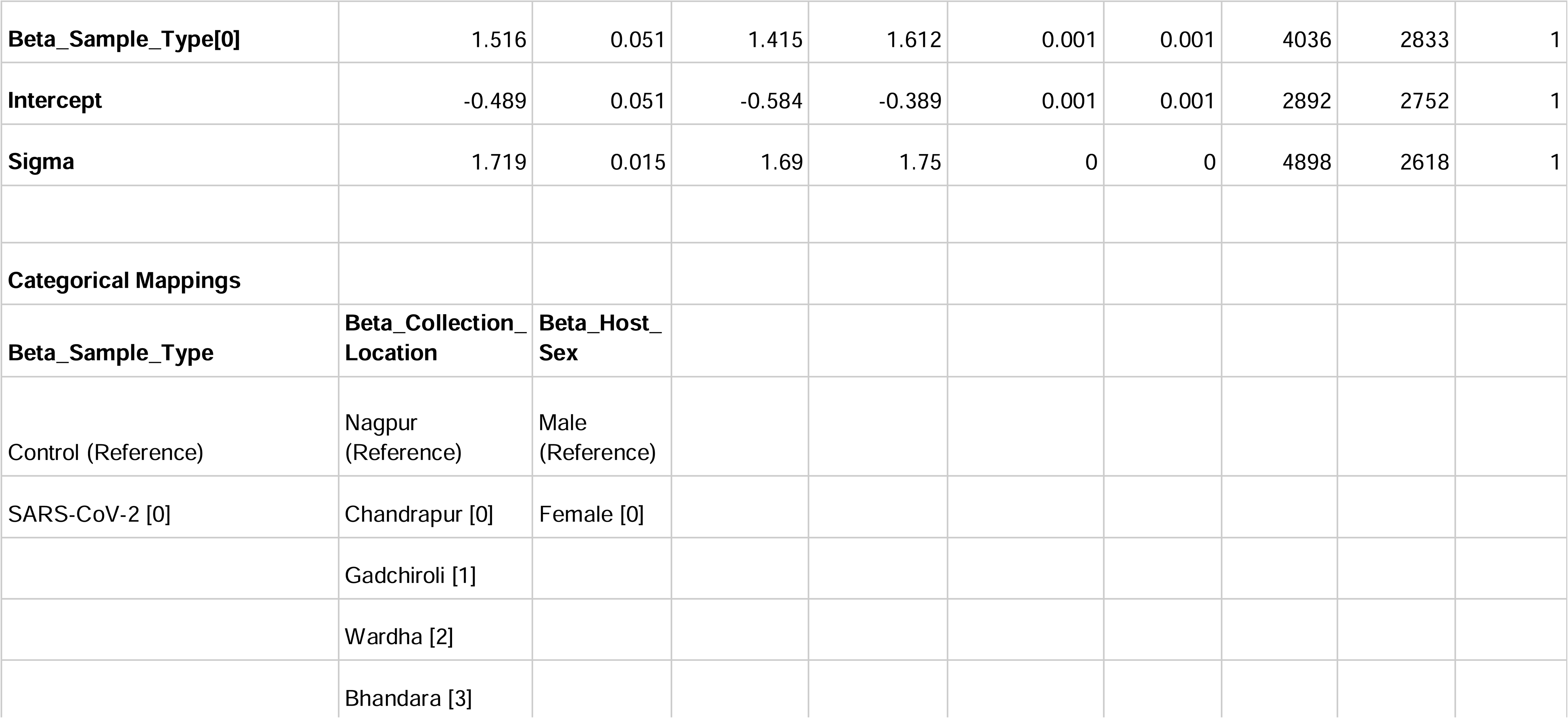
Summary of Bayesian regression model results for the effect of age, location, sex, and sample type on AMR gene abundance. Along with categorical mappings. Summary metrics include mean effects, standard deviation (SD), highest density intervals (HDI), Monte Carlo standard errors (MCSE), effective sample sizes (ESS), and convergence diagnostic (R-hat) to evaluate model reliability and convergence.

### 3.7. Antimicrobial Resistance Genes and Their Associated Drug Classes

The significant resistance genes as per volcano plot threshold (log2FC > 1 and Mann-Whitney p-values < 0.05) were analysed for their associated drug classes. The CARD database was used to attribute drug classes to gene families. The data includes 38 drug classes and 60 unique resistance genes. Some drug classes were linked to multiple genes, such as aminoglycoside antibiotics, which include AAC(3)-IIc, AAC(3)-IIe, ANT(2’’)-Ia, ANT(4’)-Ib, APH(3’’)-Ib, APH(6)-Id, and acrD. In contrast, others, like fluoroquinolone antibiotics and macrolide antibiotics, also have several resistance genes. Fewer genes represent drug classes such as mupirocin-like antibiotics and peptide antibiotics. Genes like TolC and MexB contribute to resistance within specific classes. **(Table 2)**

**Table 2:**
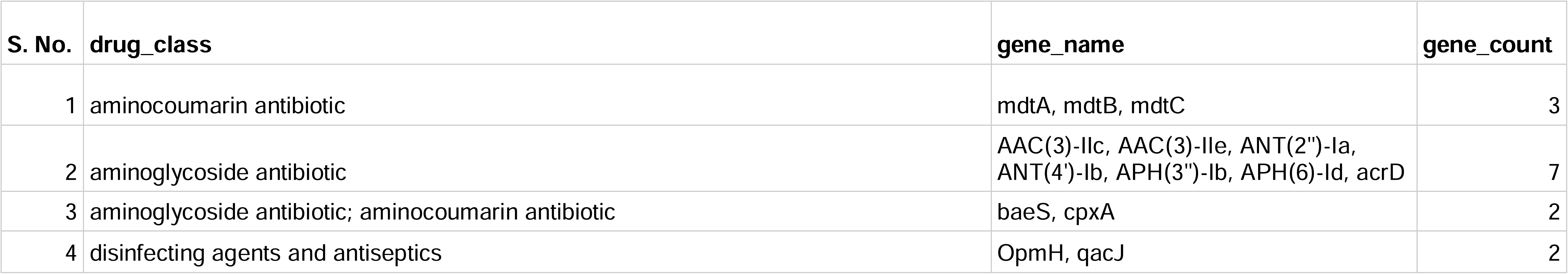
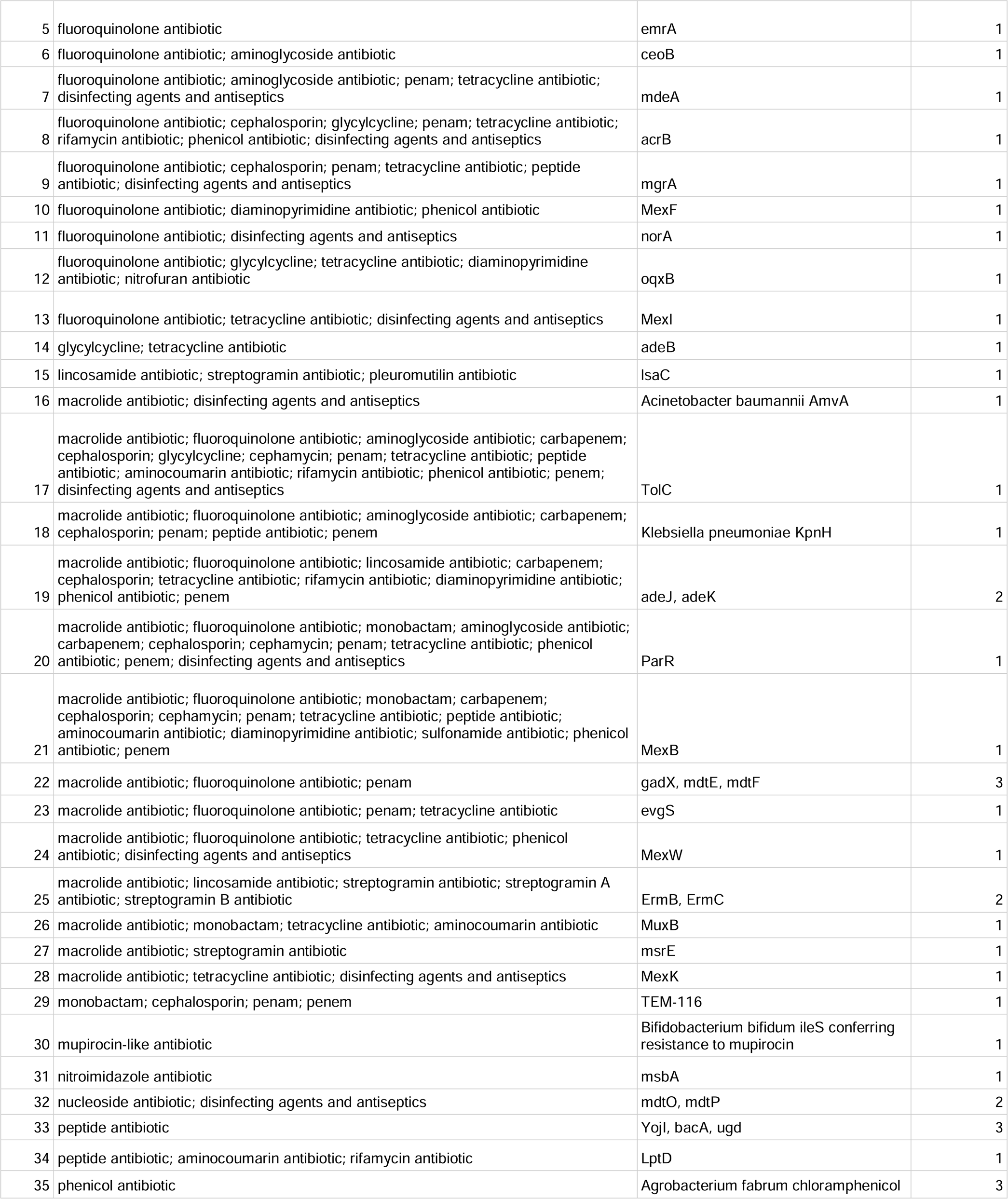
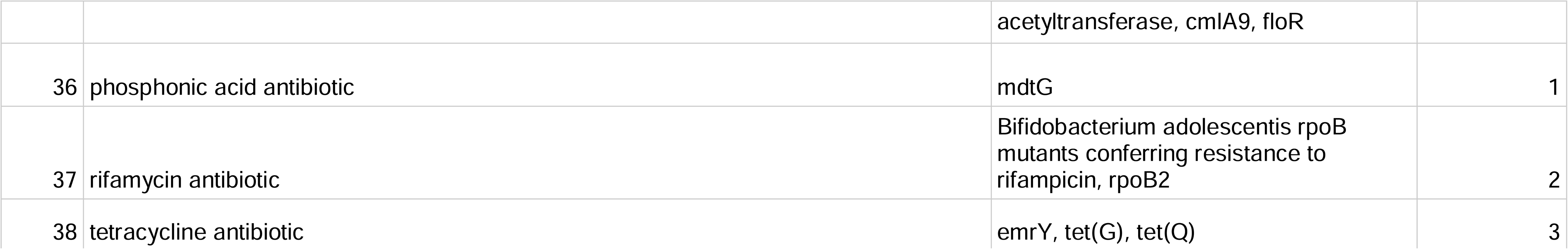
Significant AMR genes and their associated drug classes, highlighting genes contributing to resistance across multiple drug classes.

## 4. Discussion

Multiple studies around the world have investigated the compositional dynamics of URT microbiome in SARS-CoV-2 infected individuals. These studies primarily focused on explaining the relationships between the URT microbiome composition and factors like disease severity, risk of developing secondary infections and utility of URT microbiome as a marker to predict disease outcomes.^24,25,26^ It is now widely agreed upon that the SARS-CoV-2 infection have a role in altering the URT microbiome. However, the impact of infection on the AMR dynamics in the URT needs to be further looked into. Understanding the changes in AMR profiles in context with SARS-CoV-2 infection is significant for preventing secondary bacterial infection involving the lower respiratory tract and lungs. Understanding AMR dynamics could also be instrumental in devising effective therapeutics and management strategies for COVID-19 patients.

Stefanini et al. (2021)^27^ (Hoque et al., 2021)^28^ reported that SARS-CoV-2 infection is associated with higher diversity and increased abundance of AMR genes, suggesting a more complex microbial ecosystem or dysbiosis. The findings of our study also suggest that SARS-CoV-2 infection could be linked with a higher diversity and abundance of AMR genes. The sequencing results indicated a substantial number of reads generated for both groups. However, filtering for human reads significantly reduced microbial reads, particularly in the control group. This reduction of microbial reads may indicate a less abundant microbial community in the URT. The higher number of microbial reads in the SARS-CoV-2 samples indicates towards a relatively complex microbial ecosystem in the URT or a potential dysbiosis of the host microbiome due to the viral infection.

The Chao1 index demonstrated that the SARS-CoV-2 group had a greater richness of AMR genes than the control group. This finding converges with previous studies indicating that viral infections can influence the composition and diversity in the context of gut^29^ and URT^30^ microbiomes. The non-significant differences in the Shannon and Simpson indices suggest that the SARS-CoV-2 group has a diverse resistome, but the evenness of these distributions is similar to that of the control group. This finding indicates that the higher AMR richness in the SARS-CoV-2 group is not due to the skewed abundance of a few AMR genes.

Abundance of the antimicrobial resistance (AMR) genes substantially differed between SARS-COV-2 infected patients and the control group. PCA and PCoA analyses revealed distinct clustering between these two groups. Control samples exhibited lower variability in clustering, which suggests the presence of a stable and uniform resistome. Meanwhile, the SARS-CoV-2 group displayed higher variability, suggesting an active alteration of the microbial communities within the infected patients due to secondary infections, antibiotic therapies, or changes in the microbiome due to the infection.

Bray-Curtis dissimilarity further indicated the separation between the two groups. The heatmap shows the significant impact of SARS-CoV-2 infection on microbial community composition and resistome profiles. The SARS-CoV-2 group exhibited a significantly higher abundance of critical AMR genes, such as mecA, blaOXA-48, and blaNDM-1. These genes confer resistance to key antibiotics like beta-lactams and carbapenems. Elevated abundance of these critical AMR genes raises concerns about potential multidrug-resistant secondary bacterial infections in COVID-19 patients.

Pathogen-specific AMR analysis using CARD revealed stronger associations of these AMR genes with pathogens such as *Klebsiella pneumoniae*, *Escherichia coli*, and *Staphylococcus aureus* in the URT of SARS-CoV-2 patients. This finding underscores the increased risk of secondary infections, exacerbated by prolonged hospital stays, invasive procedures like intubation, or immune dysregulation during SARS-CoV-2 infections. Meanwhile, in the control group, AMR genes were primarily linked to *Acinetobacter baumannii* and *Pseudomonas aeruginosa* but at lower abundances than the SARS-CoV-2 group.

Bayesian regression analysis showed that SARS-CoV-2 infection is an important factor contributing towards increased AMR gene abundance; this finding aligns with previous studies where SARS-CoV-2 patients showed elevated AMR profiles^31,32,33^. Intriguingly, the analysis identified higher AMR gene abundance in females than in males. This finding still leaves the scope for further validation through large-scale studies that incorporate the participants’ behavioural and socio-economic variables. It is also important to highlight that in India, females tend to self-medicate more as compared to males^34,35^. The observed higher prevalence of AMR in females could also be due to this behavioural pattern. In this study we did not observe significant age-associated variations in AMR abundance and diversity. Our findings diverge from previous studies where age is identified as a significant factor contributing to AMR diversity^36,37^. The age-related paradigms may further be explored in larger longitudinal studies specifically designed to ascertain these effects.

The resistome analysis also revealed broad resistance across multiple antibiotic classes, including aminoglycosides, macrolides, and fluoroquinolones. These findings highlight the complexities of managing secondary bacterial infections in COVID-19 patients, particularly in the context of multidrug-resistant pathogens.

## Conclusion

The findings of this study highlight the need for monitoring the dynamics of AMR in SARS-CoV-2 patients. In clinical settings, secondary bacterial infections and multidrug-resistant pathogens make pandemics like COVID-19 difficult to manage. Policymakers should look forward to incorporate AMR surveillance into public health workflows. Sufficient resources must be allocated towards large-scale AMR surveillance in a populous and developing country like India. Collaboration among healthcare providers, researchers, and public health authorities is crucial to safeguarding the efficacy of antibiotic treatments and ensuring better patient outcomes.

## Limitations of the Study

The cross-sectional study design limits the ability to understand temporal changes in AMR gene profiles. Additionally, the potential impacts of antibiotic usage on AMR profiles could not be assessed in this study, as this is a retrospective study and antibiotic usage data were not collected when the samples were originally obtained for SARS-CoV-2 genome surveillance under the INSACOG mandate.

## Data availability statement

The data is available as supplementary data with the file name “**combined_supplementary_data**” Any additional data, if required, will be made available on request by the authors.

## Author declaration

The authors assure that the research has followed all ethical guidelines and received approvals from the Institutional Ethics Committee for Research on Human Subjects (IEC) of CSIR-NEERI, Nagpur-20, India. Necessary consent from patients/participants has been obtained, and relevant institutional documentation has been archived.

## Confidentiality declaration

Sample IDs (23G214-5G-264_S1 to 23G214-5G-311_S48 and 24D214-5G_387_S45 to 24D214-5G_434_S91) are masked IDs and cannot be traced to participant details. The precise age of the participants is masked, and non-overlapping age ranges were used.

## Author contribution statement

SST and KK have contributed equally to the conceptualization, experimentation, and data analysis of this study.

## Acknowledgement

The authors are thankful to CSIR-NEERI for providing funds under project OLP-57 (March 2023-April 2024) for conducting this study. This manuscript has obtained the approval of the Knowledge Resource Center (KRC) publication committee of CSIR-NEERI **(KRC No.: CSIR-NEERI/KRC/2024/NOV/EEPM/1) Date: 14-11-2024**

